# Prevalence of Undiagnosed Hypertension Among Adult Displaced Individuals in Baidoa Camps, Somalia

**DOI:** 10.1101/2024.03.22.24304736

**Authors:** Mohamed Jayte

## Abstract

**Introduction:** Hypertension may be prevalent among internally displaced people who typically do not engage in sedentary activities throughout the day. However, the prevalence of hypertension in this population remains poorly understood. In this study, we aimed to determine the prevalence of undiagnosed hypertension and associated factors among adults living in internally displaced people (IDP) camps around Baidoa.

**Methods:** A cross-sectional study was conducted, recruiting displaced persons in camps aged 18 years or older without a prior diagnosis of hypertension or current use of anti-hypertensive therapy. A standardized questionnaire was administered, and body mass index (BMI) was calculated. Hypertension was defined as two consistent measurements of systolic blood pressure ≥140mmHg and/or diastolic blood pressure ≥90mmHg, taken 4 hours apart. Multivariable logistic regression analysis was performed to identify factors independently associated with undiagnosed hypertension. Statistical significance was set at p<0.05.

**Results:** A total of 240 participants were enrolled, with a mean age of 39.4 ± 12.8 years. The majority were female (83.3%, n=199) and urban dwellers (88.3%, n=212). The prevalence of undiagnosed hypertension was 16.7% (n=40). Of the participants with hypertension, 40% (n=16) were younger than 40 years. Factors associated with undiagnosed hypertension included age >50 years (adjusted odds ratio (aOR): 7.0, 95% confidence interval (CI): 1.9 to 25.6, p=0.003), male gender (aOR: 4.2, 95% CI: 1.5 to 11.1, p=0.005), tobacco consumption (aOR: 2.6, 95% CI: 1.1 to 6.0, p=0.021), and being overweight (aOR: 3.6, 95% CI: 1.5 to 8.8, p=0.005).

**Conclusion:** Approximately one in six adult IDPs living in camps around Baidoa had undiagnosed hypertension, with a disproportionately high burden among those younger than 40 years. Further larger multi-centric studies are warranted to validate these findings.

## Introduction

Between the years 1975 and 2015, the prevalence of hypertension among adults surged from approximately 594 million cases to surpass a billion cases worldwide ^1^. Hypertension, alongside other non-communicable diseases (NCDs), stands as a primary contributor to premature mortality in both affluent and impoverished nations ^2^ NCDs alone are responsible for more than 70% of annual deaths globally, exceeding 41 million cases, with a significant portion—between 75% and 80%—of these fatalities occurring in low– and middle-income countries (LMICs)^3^ (3). Recent findings from the World Health Organization reveal that approximately 27% of adults in sub-Saharan Africa exhibit hypertension, surpassing the global average of 22% ^3^.

Hypertension exacts a toll of roughly 7% of the global burden of disability-adjusted life years, and it contributes to nearly 45% of cardiovascular morbidity and mortality worldwide ^4^. This condition is implicated in around 9.4 million fatalities, accounting for approximately 17% of all global deaths. Over the past few decades, the burden of hypertension has surged exponentially)^1^. Despite this, a considerable proportion of individuals with hypertension remain undiagnosed, untreated, or inadequately treated, thereby heightening the risk of hypertension-related complications, including stroke and myocardial infarction ^5^.

In Africa, the estimated prevalence of hypertension has steadily climbed from 54.6 million in 1990 to 92.3 million in 2000—a 70% increase— and further to 130.2 million in 2010, marking a 41% rise from 2000. Projections indicate that this figure will soar to 216.8 million by 2030, constituting a 66% rise from 2010 ^6^. In sub-Saharan Africa specifically, the total number of individuals with hypertension was estimated at 75 million in 2008 and is anticipated to reach 125.5 million by 2025 ^7^. Despite this high burden, over three-quarters of cases in Africa are unaware of their hypertension status ^8^.

In Somalia, according to the Findings from the Somali Health and Demographic Survey 2018-2019, the overall prevalence of hypertension was 26%^9^. This study also found out that the prevalence of hypertension is high in Advanced age, female gender, urban residence, and elevated wealth status were associated with higher likelihood of having a chronic disease; therefore, early identification of subjects with undiagnosed hypertension may prevent or reduce progression of many of its serious complications. Therefore, we aimed to determine the prevalence and factors associated with undiagnosed hypertension among adult Displaced People in Camps in Baidoa, Somalia.

### Study Design and Setting

I conducted a cross-sectional study among adult internally displaced persons (IDPs) residing in camps situated within a 10 km radius of Baidoa town, South west state, Somalia. The camps reside approximately 5000 internally displaced persons (IDPs). Notably, the camps lack a health wellness facility, and all medical services are accessed from nearby health centers or Bay Regional Referral Hospital, situated approximately 1 kilometer away from the camps.

#### Study Population

Adults aged 18 years and above living in the selected camps were invited to participate through home visits and announcements at community centers. Exclusion criteria were pregnancy, physical disabilities that impair measurement of blood pressure, and severe mental illness. Participants were enrolled consecutively until the required sample size per camp was reached, based on probability proportional to size sampling.

#### Sampling Technique and Procedure

The study employed a systematic sampling approach, ensuring that each individual within the population had an equal opportunity for selection. However, participants with pre-existing diagnoses of hypertension, individuals undergoing anti-hypertensive treatment, those with known heart conditions, pregnant women, and individuals under the age of 18 were systematically excluded from the study. Subsequently, the next eligible participant in the sequence was enrolled. This method facilitated the acquisition of samples that accurately represented the population while concurrently mitigating potential biases and optimizing time efficiency.

#### Sample Size Determination

The sample size for this study was determined utilizing the Kish–Leslie formula (1965). Considering a 95% confidence level (corresponding to a standard value of 1.96), a presumed prevalence of undiagnosed hypertension set at 50% due to the absence of prior studies, a maximum acceptable marginal error of 5% (0.05), and a statistical power of 80%, the calculated minimum sample size was 385 participants. Accounting for the finite population factor due to the estimated total population of 5000 individuals across all camps in Baidoa town, a final sample size of 240 participants was determined, accommodating a 10% allowance for non-response.

#### Data Collection

A structured questionnaire, translated into Somali, was administered in person to gather socio-demographic data, encompassing age, gender, educational attainment, marital status, tobacco and khat usage, duration of displacement, and family history of hypertension. Height, weight, and blood pressure measurements were acquired utilizing standardized protocols. Blood pressure readings were obtained in triplicate using a digital sphygmomanometer following a 5-minute period of rest in a seated position, with the average of the final two readings utilized for analysis.

Weights of the participants were measured employing a calibrated stand-on weighing scale. Each participant stood upright on the scale without footwear and with no additional weight in their hands or pockets, and their weight was subsequently recorded. Heights were determined using a height board, with participants standing barefoot on the height meter in an upright position, head forward, and arms hanging naturally at their sides. The maximum height was measured and recorded in meters. Body Mass Index (BMI) was then computed by dividing the weight in kilograms by the square of the height in meters to ascertain whether the respondent fell into the obese category.

### Operational Definitions

- Obesity: Defined as a Body Mass Index (BMI) greater than 30 kg/m^2.
- Overweight: Defined as a Body Mass Index (BMI) falling between 25 and 29.9 kg/m^2.
- Hypertension: Defined as a systolic blood pressure (SBP) measurement of at least 140 mmHg and/or a diastolic blood pressure (DBP) measurement of at least 90 mmHg, with readings taken at least 4 hours apart.

### Statistical analysis

The data underwent initial cleaning and preparation using Microsoft Excel 2016 before being imported into STATA Software Version 17.0 for analysis. Continuous variables, such as age and blood pressure, were re-categorized into categorical outcomes following established guidelines. Subsequently, a bi-variable logistic regression analysis was performed to examine the association between undiagnosed hypertension and sociodemographic characteristics, as well as other risk factors. The results were presented as crude odds ratios (cOR) along with their respective 95% confidence intervals (CI) and p-values.

Independent variables demonstrating some evidence of association with the dependent variable at the bi-variable analysis level (p < 0.2) were incorporated into the multi-variable logistic regression model, alongside known risk factors of hypertension with borderline outcomes. Stepwise logistic regression analysis was then conducted, adjusting for all recognized confounders. The outcomes of the multi-variable logistic regression are delineated as adjusted odds ratios (aOR) accompanied by their respective 95% CI and p-values. A significance level of p < 0.05 was considered indicative of a robust association between the independent and dependent variables.

Variables with fewer than five outcomes were excluded from the multi-variable regression due to data sparseness, and collinearity among all variables in the model was assessed. Collinearity was deemed present if the variables exhibited a variance inflation factor (VIF) exceeding 3.

In this study, a total of 240 respondents were enrolled. The mean age of all study participants was 39.4 ± 12.8 years. The majority of participants were female, constituting 83.3% (n=199), and the predominant marital status among participants was married, accounting for 63.8% (n=153). Regarding residential distribution, 11.67% of participants resided in rural areas, while the majority, 88.33%, lived in urban areas (Table 1).

**Table 1.**
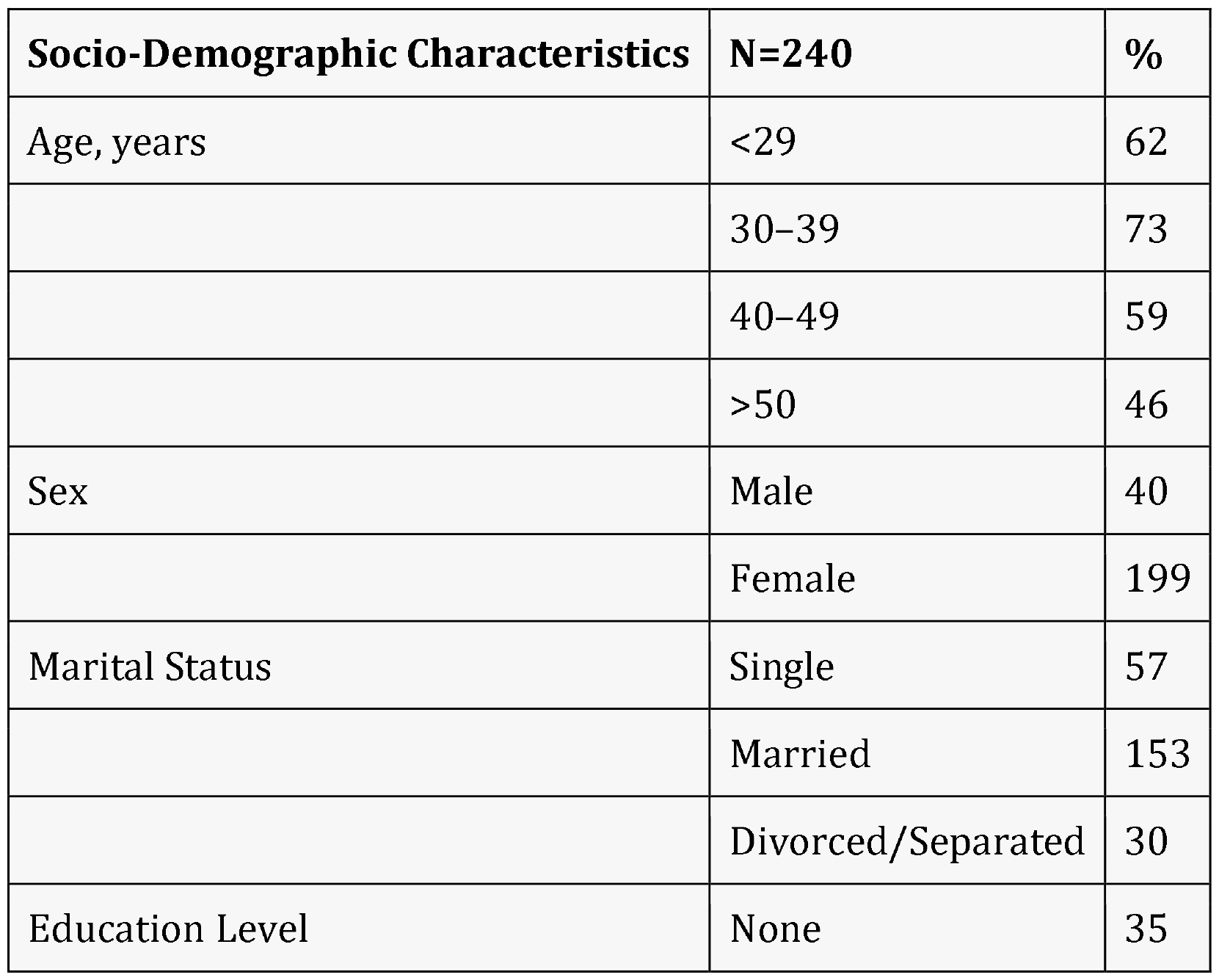
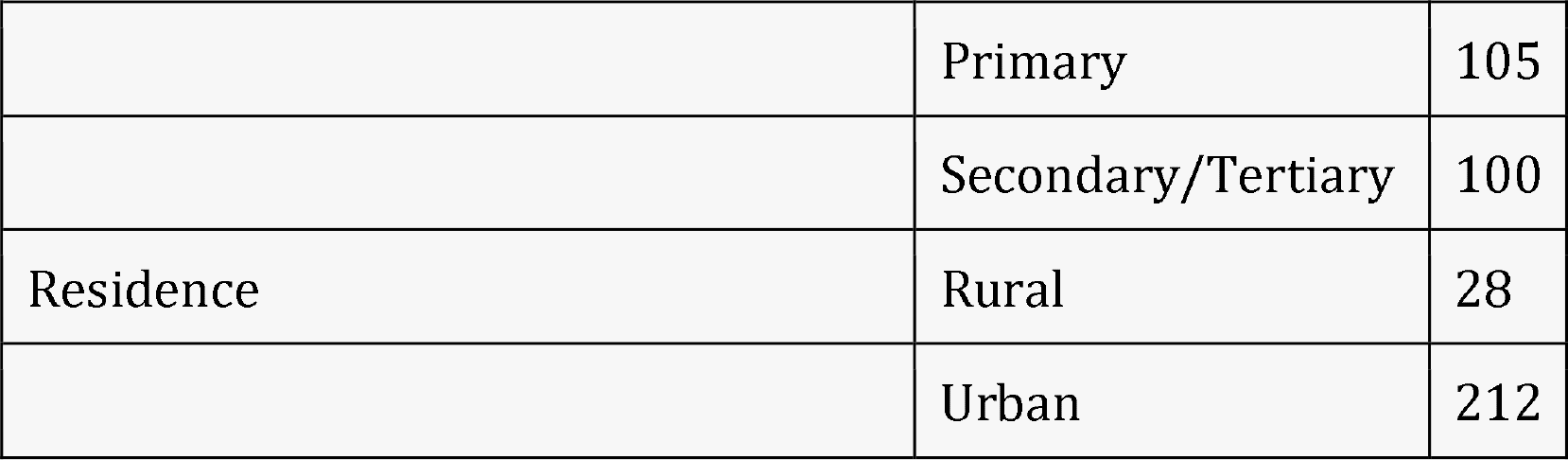
Demographic Characteristics of the Study Participants.

#### Behavioral and Socio-Economic Characteristics of the Participants

A substantial proportion, 57.5% (n=138), of the study participants reported a larger household size. Additionally, 30.8% (n=74) of participants engaged in agriculture alongside market sales. The majority, comprising 75.4% (n=181) of participants, reported no history of smokeless tobacco consumption, while only 0.8% (n=2) reported smoking cigarettes. Positive family history of hypertension was documented for 28.3% (n=68) of participants, whereas 4.2% (n=10) reported a family history of diabetes mellitus (Table 2)

**Table 2.** Behavioral and Socio-Economic Characteristics of the Participants.

**Behavioral and Socio-Economic Characteristics of the Participants**
| **Variable** | **Category** | **Frequency** | **Percent** |
| --- | --- | --- | --- |
| Household size | ≤5 | 102 | 42.5 |
|  | ≥6 | 138 | 57.5 |
| Other occupation | None | 149 | 62.08 |
|  | Agriculture | 74 | 30.83 |
|  | Others | 17 | 7.08 |
| smokeless tobacco | No | 181 | 75.42 |
|  | Yes | 59 | 24.58 |
| Duration usage of smokeless tobacco | Never | 181 | 75.42 |
|  | <1 year | 40 | 16.67 |

| Variable | Category | Frequency | Percent |
| --- | --- | --- | --- |
|  | >1 year | 19 | 7.92 |
| smokeless tobacco quantity | Never | 181 | 75.42 |
|  | 1 box | 34 | 14.17 |
|  | >1 box | 25 | 10.42 |
| Smoking history | No | 238 | 99.17 |
|  | Yes | 2 | 0.83 |
| Smoking duration | Never | 238 | 99.17 |
|  | <1 year | 2 | 0.83 |
|  | >1 year | 0 | 0 |
| Daily cigarettes smoked | 0 | 238 | 99.17 |
|  | ≥10 sticks | 2 | 0.83 |
| Physical activity | No | 99 | 41.25 |
|  | Yes | 141 | 58.75 |
| Family history of hypertension | No | 172 | 71.67 |
|  | Yes | 68 | 28.33 |
| Family history of diabetes mellitus | No | 230 | 95.83 |
|  | Yes | 10 | 4.17 |

#### Body Mass Indices of the Participants

Among the 240 participants, 49.6% (n=119) exhibited a normal BMI, while 32.9% (n=79) were classified as overweight. Additionally, 17.5% (n=42) of participants fell into the obese category (table 3).

**Table 3.**

| BMI Category | Frequency (n) | Percentage |
| --- | --- | --- |
| Normal BMI | 119 | 49.6% |
| Overweight | 79 | 32.9% |
| Obese | 42 | 17.5% |

Body Mass Indices of the Participants

#### Prevalence of undiagnosed Hypertension

Overall, 40 (16.7%) participants had undiagnosed hypertension

**Figure 2.**
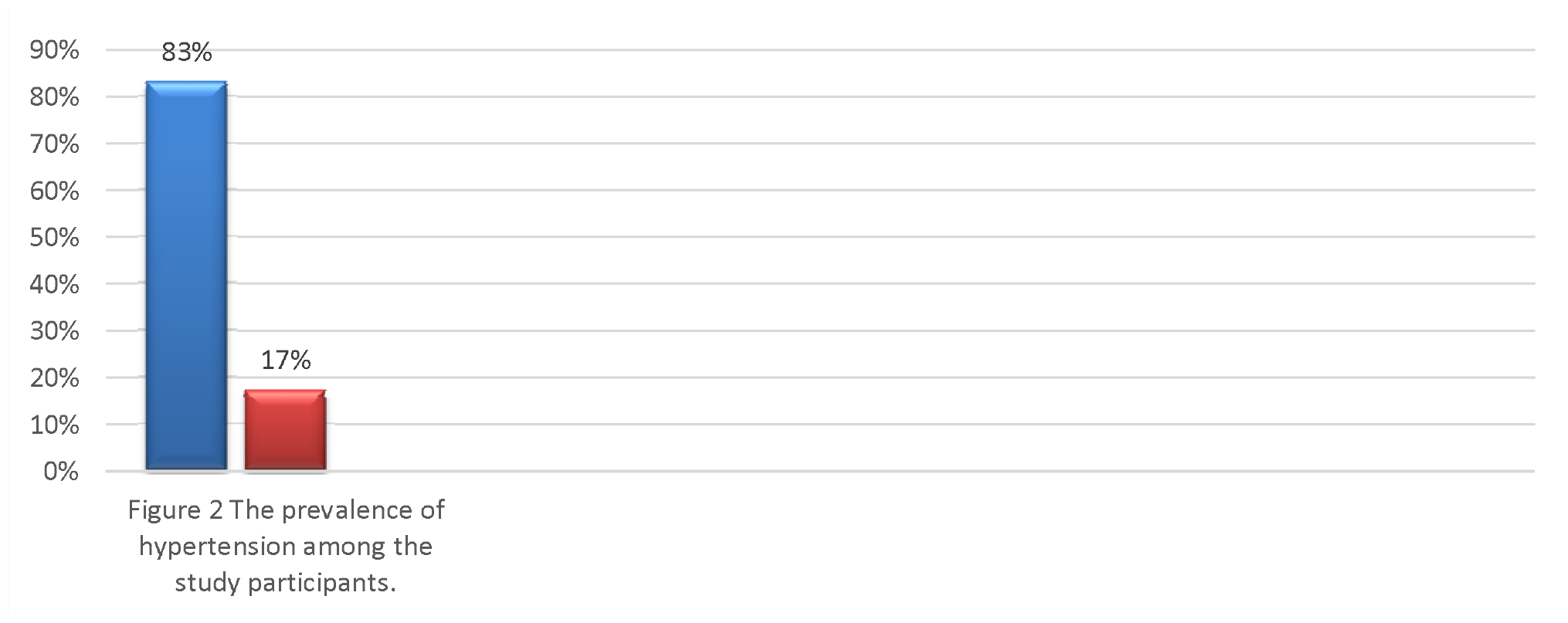
The prevalence of hypertension among the study participants.

#### Factors Associated with Undiagnosed Hypertension

The factors significantly associated with undiagnosed hypertension included age >50 years (adjusted odds ratio [aOR]: 7.0, 95% confidence interval [CI]: 1.9–25.6, p=0.003), male gender (aOR: 4.2, 95% CI: 1.5–11.1, p=0.005), alcohol use (aOR: 2.6, 95% CI: 1.1–6.0, p=0.021), and overweight status (aOR: 3.6, 95% CI: 1.5–8.8, p=0.005) (Table 3).

**Table 3.**
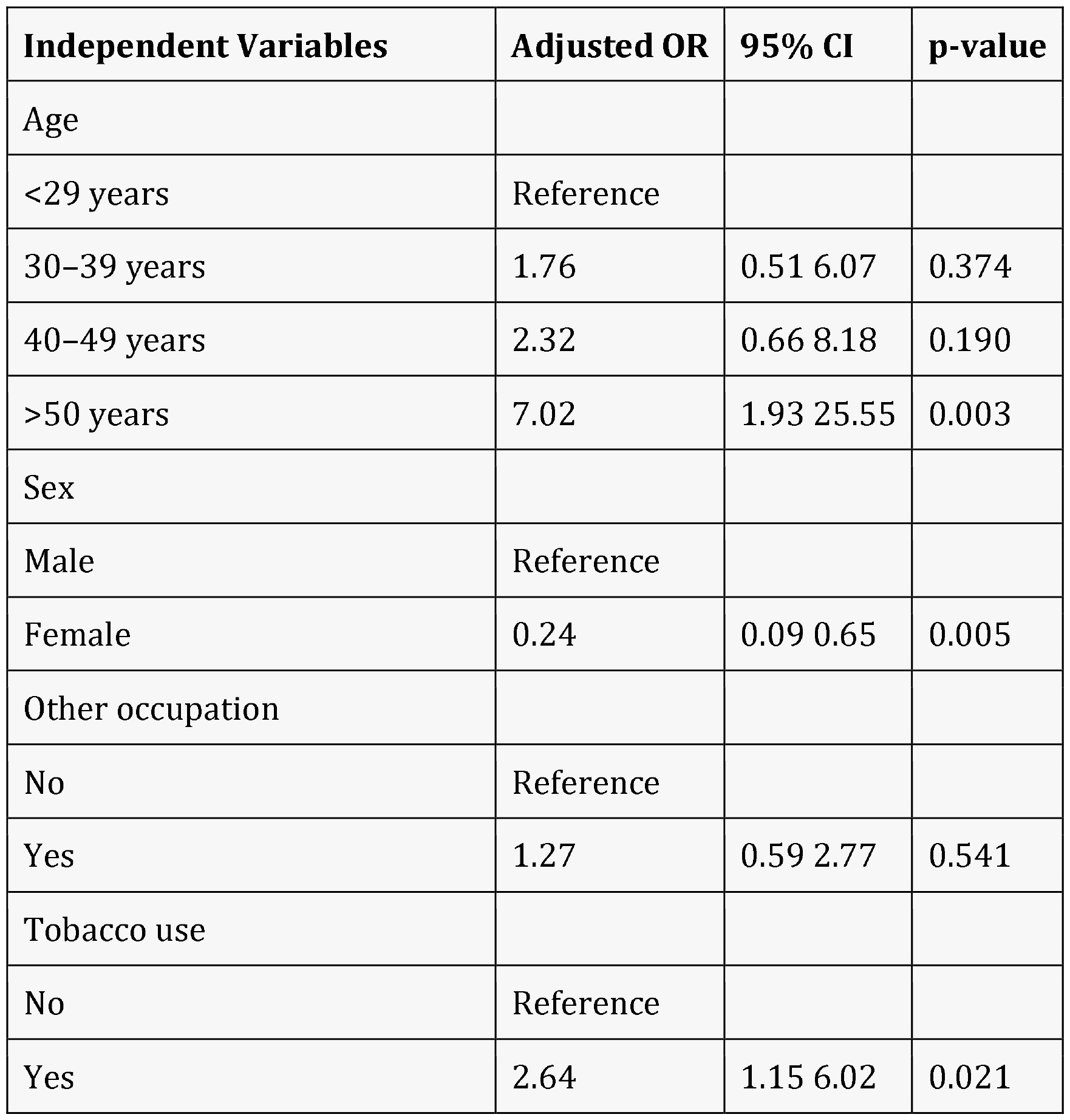
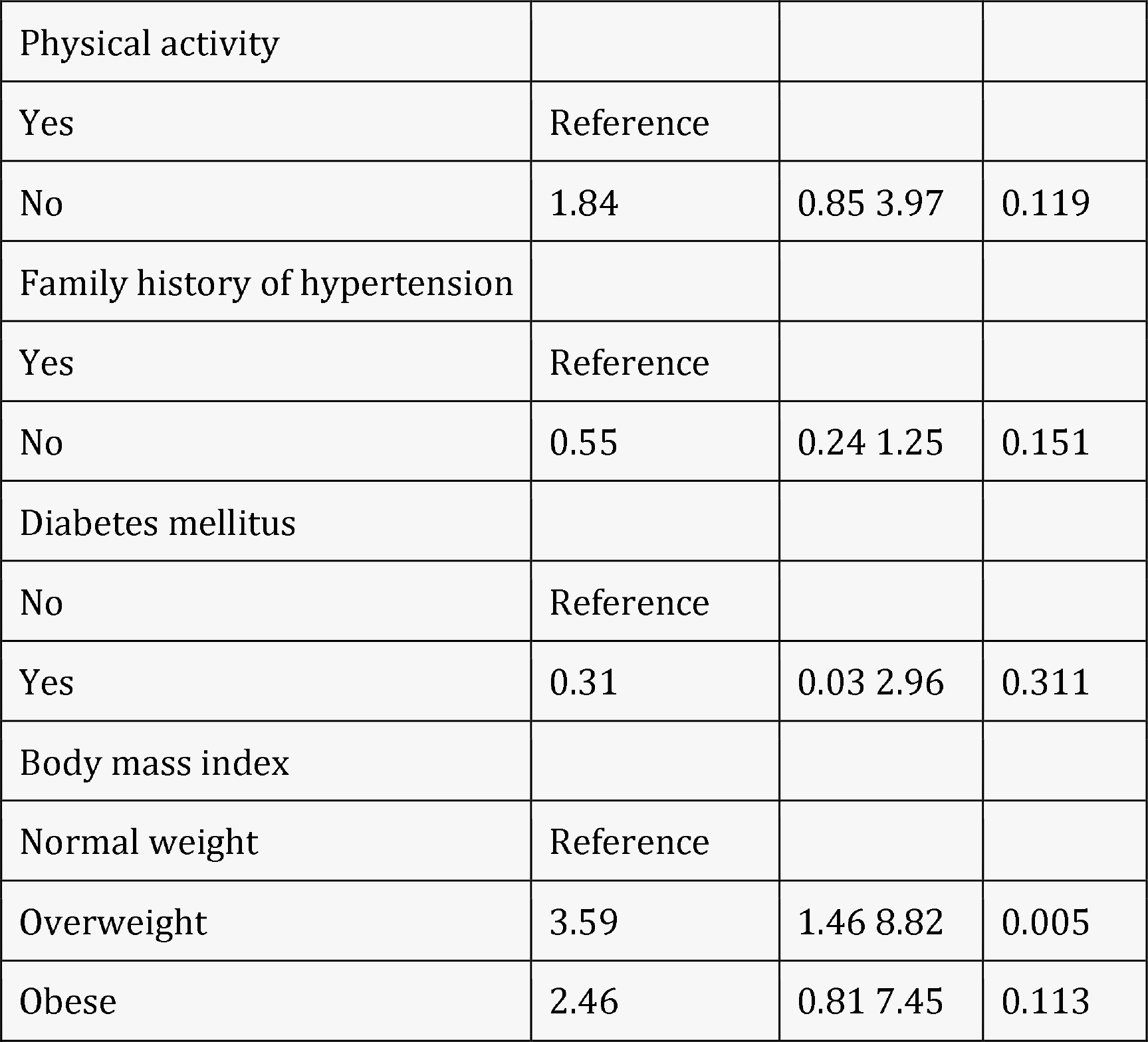
Multivariable Analysis of Factors Associated with Undiagnosed Hypertension.

| Independent Variables | Adjusted OR | 95% CI | p-value |
| --- | --- | --- | --- |
| Age |  |  |  |
| <29 years | Reference |  |  |
| 30–39 years | 1.76 | 0.51 6.07 | 0.374 |
| 40–49 years | 2.32 | 0.66 8.18 | 0.190 |
| >50 years | 7.02 | 1.93 25.55 | 0.003 |
| Sex |  |  |  |
| Male | Reference |  |  |
| Female | 0.24 | 0.09 0.65 | 0.005 |
| Other occupation |  |  |  |
| No | Reference |  |  |
| Yes | 1.27 | 0.59 2.77 | 0.541 |
| Tobacco use |  |  |  |
| No | Reference |  |  |
| Yes | 2.64 | 1.15 6.02 | 0.021 |

| **Independent Variables** | **Adjusted OR** | **95% CI** | **p-value** |
| --- | --- | --- | --- |
| Physical activity |  |  |  |
| Yes | Reference |  |  |
| No | 1.84 | 0.85 3.97 | 0.119 |
| Family history of hypertension |  |  |  |
| Yes | Reference |  |  |
| No | 0.55 | 0.24 1.25 | 0.151 |
| Diabetes mellitus |  |  |  |
| No | Reference |  |  |
| Yes | 0.31 | 0.03 2.96 | 0.311 |
| Body mass index |  |  |  |
| Normal weight | Reference |  |  |
| Overweight | 3.59 | 1.46 8.82 | 0.005 |
| Obese | 2.46 | 0.81 7.45 | 0.113 |

## Discussion

In this cross-sectional study conducted among adult internally displaced persons (IDPs) residing in camps in Baidoba, the prevalence of hypertension was approximately 17%. Our investigation revealed that older age, male gender, tobacco use, and being overweight were significantly associated with hypertension. These findings underscore a noteworthy proportion of adult internally displaced persons (IDPs) living with undiagnosed hypertension, highlighting potential gaps in awareness of blood pressure status within this population. The observed prevalence of hypertension of around 17% aligns closely with findings from a previous study conducted in Northern Somalia ^10^, which reported a hypertension prevalence of 22.6%. This consistency underscores the pervasive nature of hypertension as a silent ailment, with numerous individuals harboring the condition within the community without receiving a formal diagnosis.

The prevalence of hypertension observed in the current study aligns with previous reports from Ethiopia (12.3%) ^11^ and Uganda (16.7%) ^2^ concerning participants who were hypertensive but unaware of their condition. However, our findings diverge from a study conducted in Nakivale refugee camp, Uganda, which reported a lower prevalence of undiagnosed hypertension at 8.8%. This discrepancy may be attributed to differences in the study populations, as our investigation focused specifically on internally displaced persons, whereas the study in Nakivale refugee camp encompassed a diverse population representing individuals from various sub-Saharan African countries ^12^.

We discovered that the likelihood of hypertension escalated with age, with individuals aged between 30–39 years exhibiting 76% higher odds of hypertension compared to those under 29 years, and those aged 40–49 years displaying more than double the odds compared to the younger age group. The highest odds of hypertension were observed among participants aged over 50 years. Furthermore, our findings indicated that male gender, history of khat consumption, and overweight status were associated with increased odds of hypertension, consistent with a study conducted in Hargeisa, Northern Somalia^10^

Similarly, the aforementioned study on hypertension in Hargeisa, Somalia, revealed associations between male sex, higher BMI, and advancing age with hypertension, consistent with findings from studies conducted in sub-Saharan Africa, which identified older age as strongly associated with hypertension. These observations are supported by scientific evidence indicating that as individuals age, they are more prone to developing hypertension due to increased vascular resistance.

In light of these findings, there is a pressing need to promote healthy lifestyle practices among the elderly to mitigate the risks associated with hypertension, as evidenced by previous studies. Therefore, encouraging and implementing interventions aimed at fostering healthy behaviors among older individuals is strongly recommended.

The study further unveiled those men are at a higher risk of having undiagnosed hypertension compared to women, corroborating findings from a study conducted in a refugee camp in Uganda^12^ and results from a national survey in Kenya on the prevalence, awareness, treatment, and control, as well as determinants of hypertension Prevalence, awareness, treatment and control of hypertension and their determinants: results from a national survey ^13^. These studies demonstrated that men were less likely to be aware of their hypertension status.

This disparity in awareness between genders underscores the importance of targeted interventions aimed at improving hypertension awareness and detection among men. Efforts to enhance health education, increase access to screening services, and promote regular blood pressure monitoring among men are imperative to address this gap and facilitate timely diagnosis and management of hypertension in this papulation.

This study found out that the odds of developing undiagnosed hypertension was higher in those who use smokeless tobacco products. Our finding is also in agreement with that of a study carried out in Ethiopia^14^ which revealed that, use smokeless tobacco products among other factors like older age, were factors associated with hypertension. Our study also identified that overweight participants had nearly four times higher odds of developing undiagnosed hypertension compared to those with normal weight, followed by obese participants who had 2.5 times higher odds of developing undiagnosed hypertension. This finding is consistent with results from other studies conducted in Uganda ^2^.

This study has several limitations. Firstly, I was unable to investigate other laboratory factors such as cholesterol levels, which are known risk factors for hypertension. Additionally, while the study sample size was deemed adequate for our analysis, it is possible that certain subgroups within the population may not have been adequately represented. Furthermore, this study focused specifically on a population with limited literature, as most previous studies included both individuals who were hypertensive and those who were unaware of their blood pressure levels.

This could potentially limit the generalizability of our findings to other populations with different characteristics. Despite these limitations, our study contributes valuable insights into the prevalence and risk factors associated with undiagnosed hypertension among internally displaced persons, shedding light on an important public health concern in this population.

## Conclusion

In this study, I had observed that approximately one in six of adult IDPs living in camps around Baidoa had undiagnosed hypertension. Our findings corroborate existing evidence indicating that age, weight, sex, and Tobacco intake are significantly associated with hypertension. Notably, the highest odds of hypertension were observed among participants aged over 50 years. Given these findings, it is imperative to implement routine screening and management protocols for hypertension within this displaced population. Early detection and effective management of hypertension are crucial for reducing the burden of hypertension-related complications and improving overall health outcomes among internal displaced persons living in camps around Baidoa.

## Data Availability

All data produced in the present work are contained in the manuscript

## Acknowledgments

The author expresses his gratitude to Dr Maryan Dahir and ICR Baidoba Office for their contribution to this research.

## Data Sharing Statement

All supporting data are included within the manuscript.

## Informed Consent

Participating students provided both written and verbal consent.

## Ethical approval

The research has been approved by the Somali International University Research Board with Approval number SIU-REB-2024-001, demonstrating adherence to ethical standards.

